# A mathematically rigorous assessment of the efficiency of quarantining and contact tracing in curbing the COVID-19 epidemic

**DOI:** 10.1101/2020.05.04.20091009

**Authors:** Amaury Lambert

**Affiliations:** Laboratoire de Probabilités, Statistique & Modélisation (LPSM), Sorbonne Université, CNRS UMR8001, Université de Paris, Paris, France; Center for Interdisciplinary Research in Biology (CIRB), Collège de France, CNRS UMR7241, INSERM U1050, PSL Research University, Paris, France

## Abstract

In our model of the COVID-19 epidemic, infected individuals can be of four types, according whether they are asymptomatic (*A*) or symptomatic (*I*), and use a contact tracing mobile phone app (*Y*) or not (*N*). We denote by *f* the fraction of *A*’s, by *y* the fraction of *Y*’s and by *R*_0_ the average number of secondary infections from a random infected individual.

We investigate the effect of non-electronic interventions (voluntary isolation upon symptom onset, quarantining private contacts) and of electronic interventions (contact tracing thanks to the app), depending on the willingness to quarantine, parameterized by four cooperating probabilities.

For a given ‘effective’ *R*_0_ obtained with non-electronic interventions, we use nonnegative matrix theory and stopping line techniques to characterize mathematically the minimal fraction *y*_0_ of app users needed to curb the epidemic. We show that under a wide range of scenarios, the threshold *y*_0_ as a function of *R*_0_ rises steeply from 0 at *R*_0_ *=* 1 to prohibitively large values (of the order of 60 – 70% up) whenever *R*_0_ is above 1.3. Our results show that moderate rates of adoption of a contact tracing app can reduce *R*_0_ but are by no means sufficient to reduce it below 1 unless it is already very close to 1 thanks to non-electronic interventions.

## 1 Introduction

In this paper, we model the SARS-Cov-2 epidemic by a multitype branching process where infected individuals can be asymptomatic or symptomatic, use or not a contact tracing mobile phone app, be cooperators or defectors.

We let *f* be the fraction of asymptomatics. To fix ideas, we assume that *f* = 1/3. Current estimates of *f* range between 20% (data from the Guangdong province [1] and the Diamond Princess cruise ship [10]) and 40% (data from Japanese repatriation flights [12] and from the municipality of Vo in Italy [7]).

In the absence of mitigation measure, we assume that the average number of secondary infections from an asymptomatic individual is *R****_a_*** = 2 and is *R_i_* = 4 from a symptomatic individual, resulting in a global average ‘natural’ *R*_0_ = 3.33, which is the geometric growth rate of the infected population. This figure is precisely equal to the point estimate in France before lockdown [16] and is in line with estimates from most countries, which range between 2.2 and 3.9 in the absence of mitigation measure [8]. See Figure 1 for a cartoon representing transmissions from *A*’s (asymptomatics) and from *I*’s (symptomatics).

**Figure 1.**
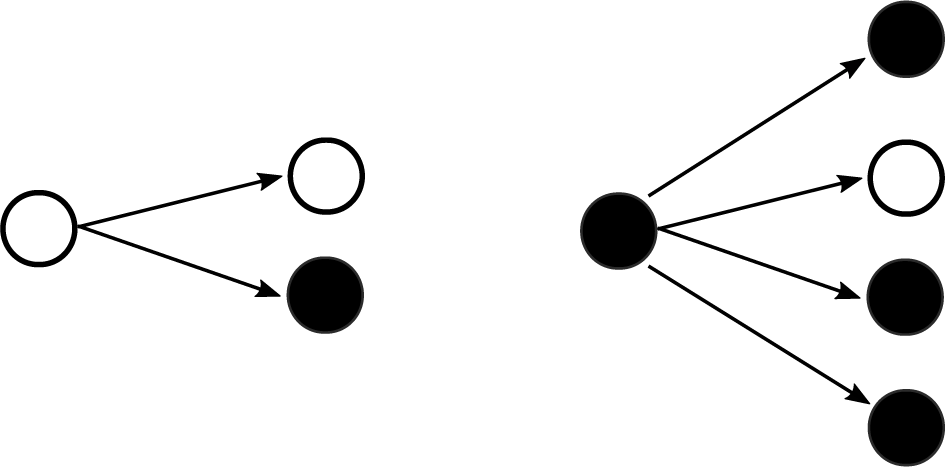
Asymptomatics (*A*) infect on average *R_a_* = 2 susceptibles and symptomatics (*I*) infect on average *R_i_* = 4 susceptibles. The overall fraction of *A*’s is *f* = 1/3. Legend: white = **A** (asymptomatic), black = **I** (symptomatic).

We explore a range of scenarios susceptible to curb the epidemic, that is, reduce *R*_0_ below 1. These scenarios are:

- Case isolation upon the appearance of symptoms;
- Additionally quarantining private contacts of symptomatics;
- Additionally quarantining physical contacts of symptomatic individuals, by means of a contact tracing app (forward tracing);
- Additionally quarantining physical contacts of physical contacts of symptomatic individuals (recursive tracing).

Let us make some preliminary observations.

First note that, except maybe in the occurrence of testings sufficiently massive to reach asymptomatics, quarantines and alerts only concern symptomatic individuals. Therefore, in the absence of mass testing, a crucial quantity is *f R_a_*, which is the growth rate of the infected population restricted to *A-to-A* transmissions (*A* denoting asymptomatics). A **necessary** condition for curbing the epidemic is then

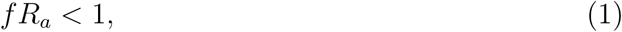

which can occur either naturally (as under our assumed default values) or by the effect of social distancing – see Section 3.

In the same vein, once *R_a_* and *R_i_* have been optimally reduced by non-electronic interventions (social distancing, case isolation), a crucial quantity is (1 − *y*)*R*_0_, where *y* is the fraction of the population not using the contact tracing app, because (1 − *y*)*R*_0_ is the growth rate of the infected population restricted to *N-to-N* transmission (*N* denoting individuals *not* using the app). A **necessary** (but certainly not sufficient, as we will see) condition for curbing the epidemic is then

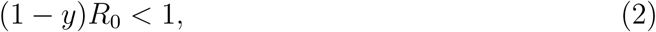

or equivalently

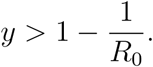

This means for example that if non-electric interventions have lowered *R*_0_ from its natural value to an effective value of 1.5, say, then the fraction of app users must be at least 1/3 for contact tracing to have a chance to work. The purpose of Sections 4 and 5 is to precisely determine the minimal value of *y* allowing contact tracing to get *R*_0_ below 1, i.e. to let the **epidemic die out, an event we will consistently denote by** (⋆).

For the sake of efficiency in these times of emergency, we summarize our results in Box 1, Theorem 1 and Figure 2.

**Figure 2.**
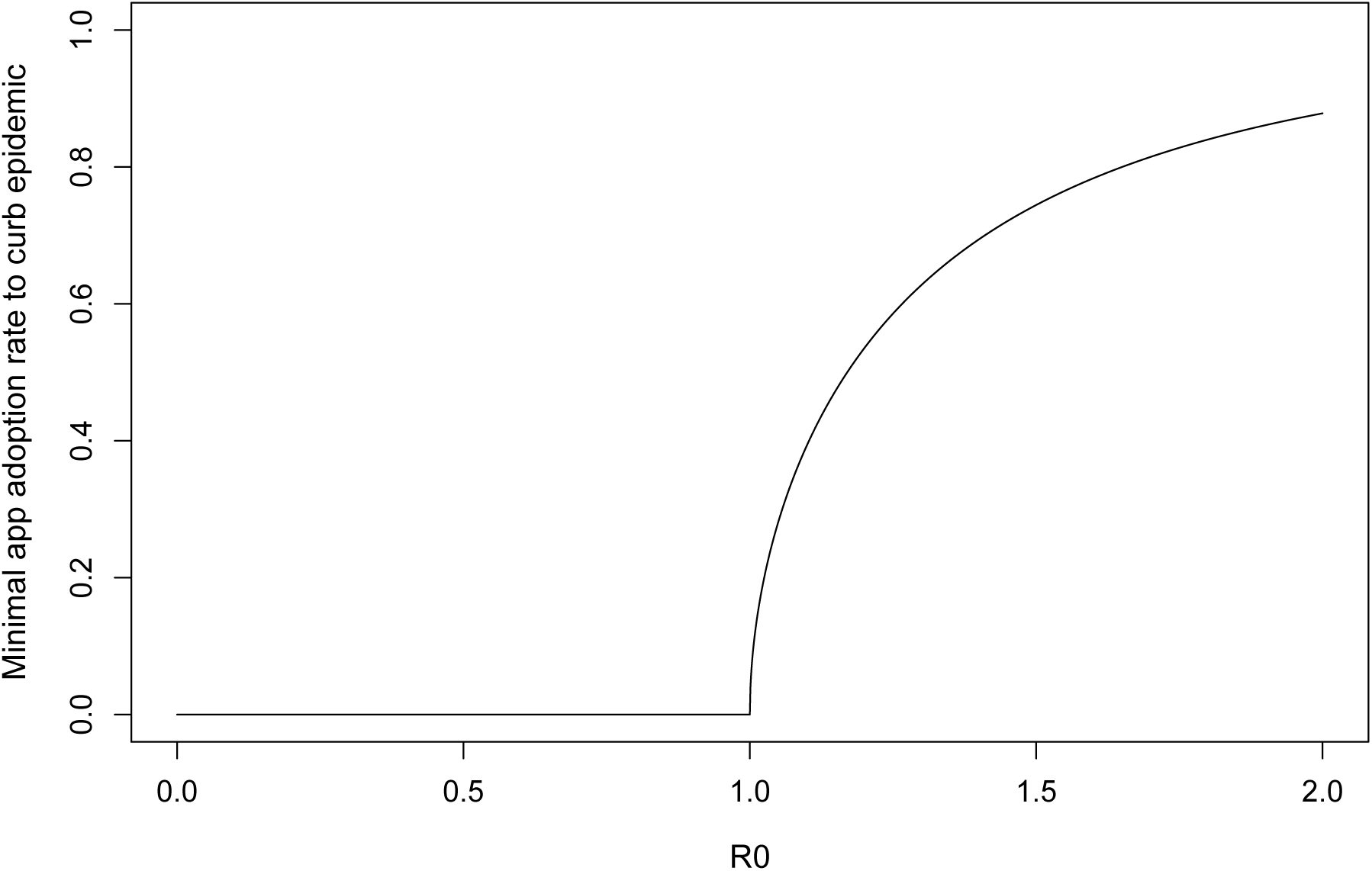
**Minimal app adoption rate yo to curb epidemic as a function of effective** *R*_0_ (i.e., obtained as a result of social distancing, case isolation and quarantining of private contacts, regardless of contact tracing), under **recursive contact tracing**. Cooperating probabilities are set equal to *q*_0_ = *q*_1_ = *q*_2_ = 1 (best case scenario). Other parameters are set to their default values: *f* = 1/3, *b* = 3/5, *kr* = 1/6, *R_a_* = 2, *R_i_* = 4. This figure is the same as top right panel of Figure 7.

#### Box 1. Summary of results

Infected individuals can be of four types, according whether they are asymptomatic (*A*) or symptomatic (*I*), and use a contact tracing mobile phone app (*Y*) or not (*N*). We investigate the effect of non-pharmaceutical interventions, depending on the fraction *f* of asymptomatics, the average number of secondary infections from asymptomatics, *R_a_*, vs from symptomatics, *R_i_*

We take default values *f* =1/3, *R_a_* = 2 and *R_i_* = 4, so the **natural** average number of secondary infections is **R**_0_ = *f R_a_* + (1 − *f*) *R_i_* = 3.33.

We first study the effect of voluntary isolation upon symptoms (thinning *R_i_* by a factor *b* = 1 − *p* + *pm*, where *p* is the cooperating probability and *m* is the fraction of transmissions made before symptoms ≈ 0.5) and of quarantining private contacts (further thinning *R_i_* by a factor 1 − *kr* that we take ≈ 0.85). Both *R_a_* and *R_i_* are additionally thinned by a factor *c* parameterizing the effect of social distancing. This unknown parameter allows us to tune the *R*_0_ of the epidemic obtained with non-pharmaceutical, non-electronic (i.e., using no contact tracing app) interventions, that we call the **effective R**_0_.

We then investigate the effect of a contact tracing app assumed to be used by a fraction *y* of the population. **For a given effective** *R*_0_, **we characterize the threshold** *y*_0_ **required to control the epidemic**, depending on how app users are willing to cooperate. We assume that an index case of type *Y I* (symptomatic, using the app) informs the app of her symptoms with cooperating probability *q*_0_, that a *Y*-individual receiving an alert self-quarantines with probability *q*_1_ or *q*_2_ depending on whether she is a physical contact of the index case (**forward tracing**) or a contact of a contact (**recursive tracing**).

Using non-negative matrix theory and stopping line techniques, we characterize *y*_0_ as the root to some smooth, explicit function (see Theorem 1 below). We show that **under a wide range of scenarios, the threshold** *y*_0_ **as a function of effective** *R*_0_ **rises steeply from 0 at** *R*_0_ = 1 **to prohibitively large values (of the order of** 60 – 70% **up) whenever the effective** *R*_0_ **is above 1.3**. Figure 2 displays a typical curve of *y*_0_ vs *R*_0_. **These figures show that moderate rates of adoption of a contact tracing app, such as currently seen in most countries, can certainly reduce** *R*_0_, **but are by no means sufficient to reduce it below 1**.

### Theorem 1

*Fix the effective average numbers of secondary infections (R_a_, R_i_, R*_0_*) obtained with non-electronic interventions (social distancing, isolation upon symptom onset, quarantining private contacts of symptomatics)*.

*Recall that* (⋆) *denotes the event that the epidemic dies out (i.e., the new R*_0_ *obtained with contact tracing is reduced below* 1).

*(i) In the case of forward tracing*,

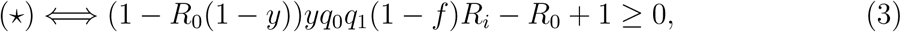

*or equivalently*

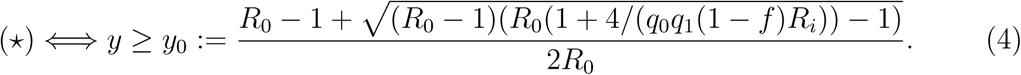

*(ii) In the case of recursive tracing*,

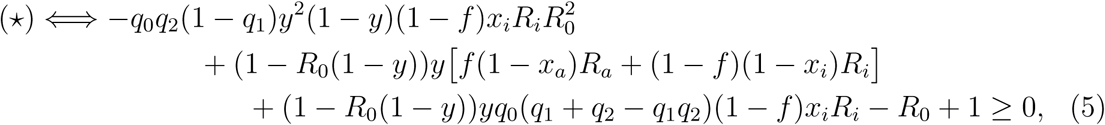

*where x_a_ =* 1 − *ℓ + ℓt_a_ and x_i_ =* 1 − *ℓ + ℓt_i_, with t_a_ (resp. t_i_) the probability that an individual of type Y, daughter of an individual of type Y A (resp. of type Y I) does not comply to any alert coming from her siblings of type Y I, given by Eq* (7) *page 15*.

Part (*i*) of the previous statement is proved page 11 and part (ii) is proved page 15 onwards (Subsection 5.3).

### Remark 1

*We recover from Eq* (3) *the fact that* y *must be at least* 1 − 1/*R*_0_ *in order to control the spread. The actual threshold y*_0_ *given in Eq* (4) *is actually much higher*.

### Remark 2

*Note that when q*_2_ = 0, *there is no additional effect of recursive tracing compared to forward tracing, and indeed Eq* (5) *becomes (because x_i_ is then equal to* 1*) exactly, as expected, the equation obtained in the case of forward tracing (Eq* (3)).

### Remark 3

*Note that when q*_1_ = 1, *the only additional effect of recursive tracing compared to forward tracing is the alert of siblings and indeed Eq* (5) *becomes in this case*

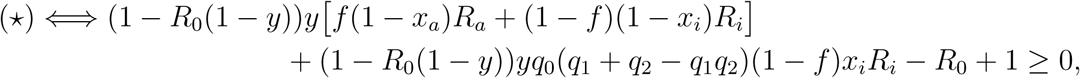

*which boils down to Eq* (3) *whenever x_a_ and x_i_* (*embodying the effect of siblings’ alerts*) *are set to* 1 *(for example by taking ℓ* = 0).

## 2 Basic modeling assumptions

We call a given infected individual a **case** or **index case**, and **secondary infections** the individuals she infects. We will also speak of **mother**/**daughter** (and not donor/receiver, because one can receive both an alert and a virus).

Now a typical index case can, **independently**:

- be **Asymptomatic** (**A**) with probability **f**, or **Symptomatic** (**I**) with probability 1 − *f*. We will take *f* = 1/3 as the default value (see Introduction). We denote the average number of secondary infections from an *A*-individual by **R**_a_ and the average number of secondary infections from an *I*-individual by **R**_i_. The mean number of secondary infections in the population is then

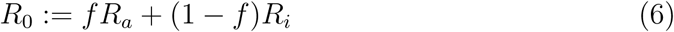
- **use a contact tracing app** (**Y** for yes) with probability **y**, or **not** (**N**) with probability 1 − *y*.
- **cooperate** or **defect**. Here, cooperating/defecting can mean several things:

–A symptomatic individual can cooperate by **self-isolating upon symptom onset**, and thus ceasing to infect other people after that time (probability of cooperating p).
–A **private contact** (work, family…) of a symptomatic individual can also cooperate by **self-quarantining after being alerted by plain talk/phone/email**, and thus ceasing to infect other people after that time (probability of cooperating **r**).
–A symptomatic individual who uses a contact tracing mobile phone app can cooperate by **entering the information in her app** as soon as she feels the first symptoms (probability of cooperating **q_0_**).
–An individual who uses the app and is alerted by her app can cooperate by **self-quarantining upon receiving the electronic alert**. We will distinguish whether the receiver of the alert has degree 1 or 2 with the original index case in the contact network (probabilities of cooperating **q_1_** and **q_2_**, respectively) – see Sections 4 and 5 for details.

### Important assumptions

We will make the following two assumptions.

- **Branching assumption**. We assume **independence of infection events** (branching assumption). This means in particular that 1) susceptibles are always in excess and that 2) the contact network is tree-like, neglecting the existence of shared contacts. This biases our predictions in two ways, because 1) we neglect the possible reduction of the effective *R*_0_ thanks to the accumulation of recovered, immune individuals; 2) we underrate the efficiency of alerting in case when transmission has occurred in clusters, but we also underrate the speed of propagation by ignoring these clusters.
- **Multiple alerts**. Note that an individual can be confronted to the decision of cooperating or defecting several times and in particular be alerted by several different sources (private vs public, single source of degree 1 vs several sources of degree 2). In contrast to standard models where individuals stick to the same strategy of cooperating or defecting, we assume here that each time an individual can **cooperate or defect, she decides to do so** with the aforementioned probabilities, **independently and independently of her past decisions to cooperate or defect**. In addition, for defection to actually occur, **an individual receiving several alerts must defect independently to each of these alerts**, modeling the multiplicative effect of multiple alerts.

### Non-electronic mitigation measures

The values of *R_a_*, *R_i_* and *R*_0_ may vary (but Eq (6) always holds) according to **four scenarios of non-electronic interventions (i.e., independent of contact tracing):**

- **No intervention**. In this case, we use the notation 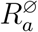, 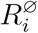 and 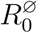. We will take as default values 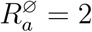 and 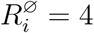, so that 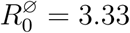.
- **Social distancing**. In this case, we use the notation 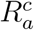, 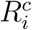, 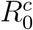. We assume that social distancing scales both 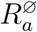 and 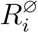 by a factor *c*, where **c** is the **reduction of infections due to social distancing**, so that

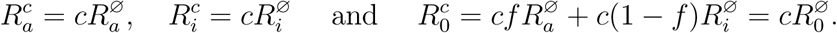
- **Additionally self-isolating upon the appearance of symptoms**. In this case, we use the notation 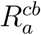, 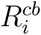 and 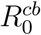. We will assume that self-quarantining scales (only) 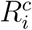 by a factor **b** = 1 − *p* + *pm*, where **m** is the **average fraction m of secondary infections made before first symptoms** or before case isolation actually starts, and **p** is the probability of cooperating, i.e., of actually isolating upon symptoms (see below), so that

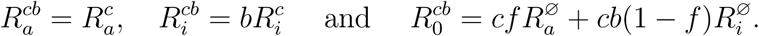
- **Additionally quarantining private contacts of symptomatics**. In this case, we use the notation 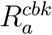, 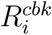 and 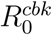. We will assume that symptomatic individuals alert their **private contacts**, who represent a **fraction k of secondary infections**, and that a fraction **r** of them does self-quarantine. See Figure 3. Then we get

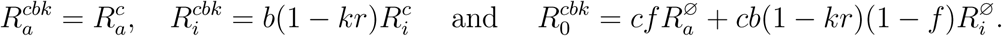

**Figure 3.**
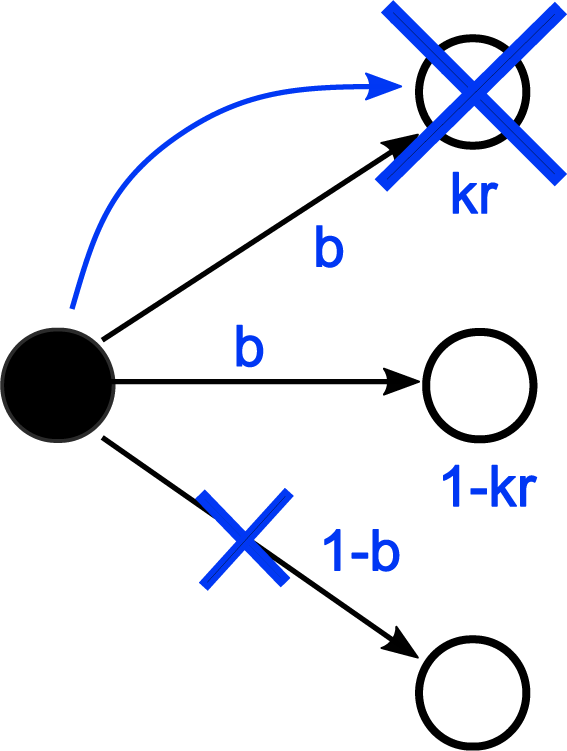
Upon symptom onset, an *I*-individual decides with probability *p* to self-isolate, resulting in removing an average fraction 1 − *b* = *p*(1 − *m*) of her daughters from the epidemic (small, blue cross). A fraction *k* of the ‘surviving’ daughters is assumed to be private contacts, who are alerted (blue arrow) and then do self-quarantine with probability *r* (large, blue cross). Legend: white = **A** (asymptomatic), black = **I** (symptomatic).

## 3 Non-electronic interventions

### 3.1 Social distancing

Social distancing scales indistinctively 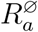 and 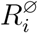 by a factor *c*, so that 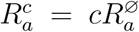, 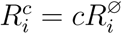 and 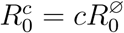.

#### Remark 4

*As mentioned in the introduction*, 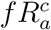 *is the growth rate of the epidemic restricted to A-to-A transmissions, that cannot be controlled in the absence of mass testings. It is then necessary for the epidemic to die out to have*

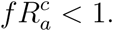

### 3.2 Self-quarantining

Assume a certain fraction **p** of symptomatic individuals **cooperate by self-quarantining** upon day *D* + *T*, where *D* is the day of onset of symptoms and *T* is the waiting time before taking action, like *T* = 0, 1 or 2. Let m be the **average fraction of the total number of secondary infections, that are already made by** day *D* + *x* from a typical symptomatic individual. For *T* = 0, *m* ≈ 0.4 [9]. We will take *m* = 0.5 as default value.

Now set

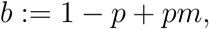

so that 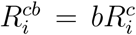 is the average number of secondary infections from an *I*-individual in this self-quarantining regime, whereas 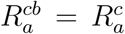 remains unchanged. Note that *b* is bounded from below by the average fraction of secondary infections transmitted during the incubation period (i.e., before first symptoms).

In this scenario, the epidemic dies out iff

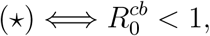

where 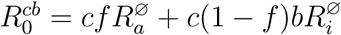.

#### Application

Here we use the default values *f* =1/3, 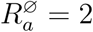 and 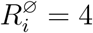.

If we assume that 100% of symptomatic individuals self-isolate upon symptoms (*p* = 1), then *b* = 1/2, and we need *c* < 1/2 to control the epidemic, which means that social distancing would have to cut down transmissions by at least 50%.

If we assume that only 50% of symptomatic individuals self-isolate upon symptoms (*p* = 1/2), then *b* = 3/4, and we need *c* < 3/8, which means that social distancing would have to cut down transmissions by at least 62.5%.

The possibility that mere social distancing does not come anywhere near these figures cannot at all be discarded. In such a situation, case isolation would not be sufficient in itself to curb the epidemic. We will take as **default value b = 3/5**, which corresponds to the **optimistic estimate of a fraction p = 4/5** of cooperators and yields 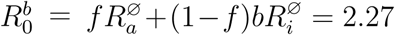. Then the average number of secondary infections in the presence of both social distancing and self-quarantining is 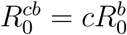, which remains larger than 1 whenever *c* > 44%.

For example, with *c* = 3/4, 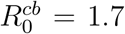, with *c* = 2/3, 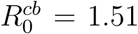 and with *c* = 1/2, 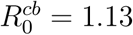.

We will now investigate (assuming *c* > 0.44) the effect of quarantining private contacts (which does not require a contact tracing app).

### Self-isolating and quarantining private contacts

Here we consider the possibility that symptomatic individuals alert, by plain talk/phone/email their **private contacts** (work, family). We denote by **k** (for ‘known’) the fraction of **secondary infections that are private contacts** and by **r** the fraction of **private contacts who are alerted and do self-quarantine**. We assume that all self-quarantining daughters are removed from the epidemic because they self-quarantine before being infectious.

In this scenario, the epidemic dies out iff

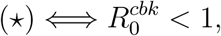

where 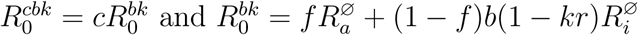

#### Application

In [14], out of a total of 7,324 well documented cases in 120 Chinese towns in January-February 2020, only 1,245 could be clustered into mini outbreaks involving 3 or more people in the same household, transport, restaurant, mall … These figures show that private contacts play a minor role in the epidemic so that in reality *k* is quite low. Note that in this study, outbreaks of 2 people were excluded from the study, most of which were spouse-to-spouse transmissions –but even if these potential transmissions are numerous, they are expected to be associated to a very low *r*. If we take *kr* = 1/6 and stick to the default values given earlier of *f* = 1/3, 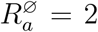, 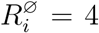 and *b* = 3/5, we get 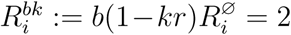, so that 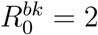, and we need *c* < 1/2 to curb the epidemic.

We will call 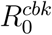 the **effective** *R*_0_, compared to the **natural** 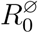. We will now assume that *c* > 1/2 and investigate for a given effective *R*_0_, whether a contact tracing app can manage to control the spread.

## 4 Forward contact tracing

Now we assume that a proportion *y* of the population uses a contact tracing mobile phone app. Such individuals are denoted **Y** (‘yes to the app’), and the others **N** (‘no to the app’).

In this subsection we consider as a first step that an alert is always of **degree 1**, that is, originates from an individual of type *YI* (using the app, symptomatic) and is only transmitted to her close physical contacts. We reserve for the next section the case of **alerts of degree 2**, that is, which originate from the index case *YI* but are conveyed through an intermediate physical contact of the case, to a contact of this contact.

Recall the probabilities of cooperation *q*_0_ and *q*_1_ defined as follows. A *Y I*-individual informs the app of her symptoms with probability *q*_0_ and a *Y*-individual alerted by a cooperating *Y I* self-quarantines with probability *q*_1_. We assume that:

- decisions to cooperate or defect that do not require the app (self-quarantining and alerting private contacts) are independent of using the app or not.
- decisions of the same individual to cooperate or defect in different situations are independent.

Let us compute the average number of secondary infections in each class, *N* or *Y*, depending on the class of the index case, *YI*, *YA*, *NI*, *NA*.

Let us start with an index case *Y I* (using the app, symptomatic) who feels her first symptoms. An individual *Y* infected by this index case:

- receives a private injunction to self quarantine with probability *k* and if this is the case, cooperates with probability *r*;
- receives independently an electronic injunction with probability *q*_0_ and if this the case, cooperates with probability *q*_1_.

Then the probability for a *Y*-individual of not being removed when infected from a *Y I* - individual is

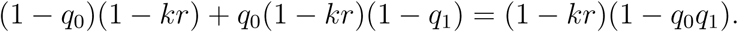

In cases when the mother is symptomatic (*I*), but either she (*NI*) or her daughter (*N*) is of type *N*, this probability is always 1 − *kr*. In cases when the mother is asymptomatic (*Y A* or *N A*), this probability is always 1, regardless of the daughter’s type (*Y* or *N*).

Now let *M* be the matrix with entries *m_y_,_y_*, *m_y_,_n_*, *m_n_,_y_*, *m_n_,_n_*, where *m_y_,_y_* denotes the average number of secondary infections of type *Y* made by a random *Y*-individual and so on and so forth. We easily get

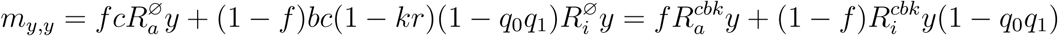

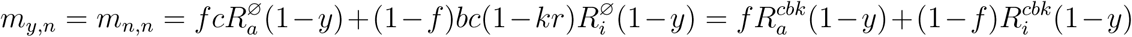

and finally

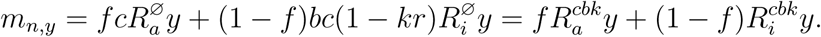

### Remark 5

*We see from the previous equations that whenever the nature and number of non-electronic interventions enforced is fixed and known, the mean numbers of secondary infections from Y/N to Y/N only depend on these interventions through* 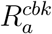 *and* 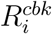*. Then from now on, we will drop superscripts and use the generic notation R_a_ and R_i_, defined as* ***effective*** *average numbers of secondary infections. The values of these two parameters will depend upon the nature and number of non-electronic interventions enforced, or more precisely on the values of c, b, k and r. Using the same default values as in the previous section yields R_a_* = *R_i_* = 2*c*.

**Figure 4.**
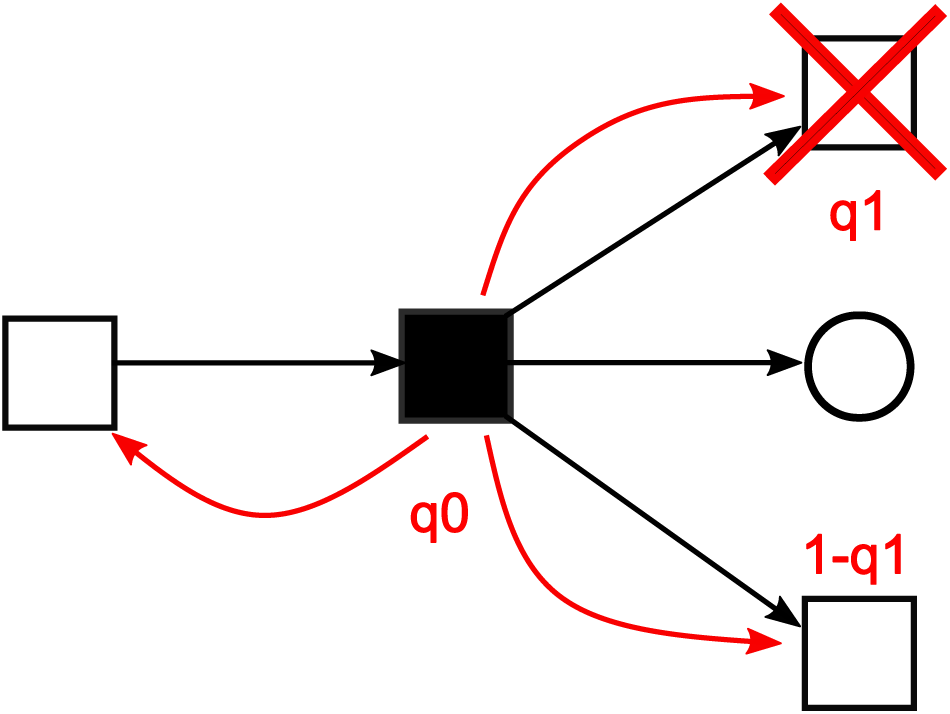
A *Y I*-individual (symptomatic, using the app) decides with probability *q*_0_ to inform the app of her symptoms, resulting in alerting (red arrows) all her physical contacts of type *Y* (among which her mother and daughters in the epidemic). Each alerted daughter does self-quarantine with probability *q*_i_ and is then removed from the epidemic (red cross). Legend: white = **A**, black = **I**, square = **Y**, circle = **N**.

We now prove Theorem 1(*i*). Using the notation 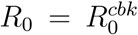, 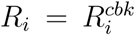 (see previous Remark) and *s_i_* = *q*_0_*q*_1_ (1 − *f*)*R_i_*, we get

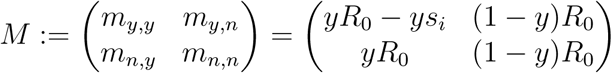

It is well known that the epidemic dies out if the leading eigenvalue of *M* is smaller than 1 [11]. Now let *Q* be the characteristic polynomial of *M*:

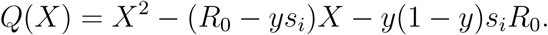

Since *Q*(0) ≤ 0, the leading eigenvalue of *M* is the unique positive root of *Q*. Also, this root is smaller than 1 if and only if *Q*(1) ≥ 0, which yieldsh

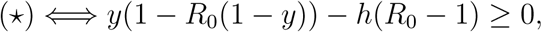

where

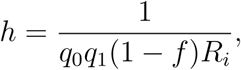

which is exactly Eq (3) of Theorem 1(i).

We define ***y*_0_** the **minimal fraction of users of the app** in order to control the spread, or **minimal app adoption rate**, by

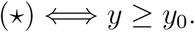

elementary calculus yields

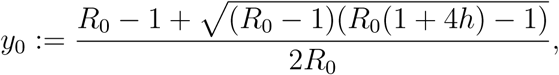

exactly as in Eq (4) of Theorem 1(i).

### Remark 6

*Note that for electronic interventions to be able to curb the epidemic, we need* y_0_ *to be actually smaller than* 1*, that is*,

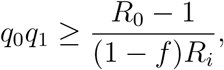

*which can from the start only hold if* (*R*_0_ − 1)/(1 − *f*)*R_i_* < 1. *This inequality is equivalent to f R_a_* < 1, *which we have already seen as Eq* (1). *Indeed, if f R_a_* > 1, *the epidemic restricted to A-to-A transmissions would be growing exponentially, with no control possible by neither electronic nor non-electronic interventions* (*in the absence of mass testing*).

### Application

The actual values of *R*_0_ and *R_i_* depend on the nature and strength of non-electronic interventions. Let us assume that social distancing is in force, parameterized by an unknown scaling factor c, in addition to case isolation and quarantining of private contacts (respectively parameterized by b and *kr*). If we stick to the default values given earlier (*f* = 1/3, 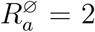, 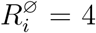, *b* = 3/5, *kr* = 1/6), which yield *R*_0_ = *R_i_* = 2*c*, we can study how *y*_0_ varies as a function of the effective *R*_0_.

If social distancing cuts down infections by 1/4, i.e., if *c* = 3/4, then the effective *R*_0_ = 3/2 and

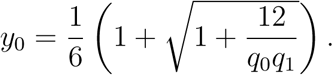

Note that *q*_0_*q*_1_ is the probability that the index case using the app does enter the information about her symptoms into the app and that the physical contact receiving the alert does self-quarantine. For the threshold *y*_0_ to be smaller than 1, we need that *q*_0_*q*_1_ ≥ 1/2. If app users are 100% reliable, that is *q*_0_*q*_1_ = 1, we get that the minimal adoption rate of the app is

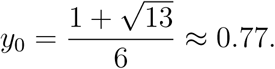

If social distancing cuts down infections by 1/3, i.e., if *c* = 2/3, then the effective *R*_0_ = 4/3 and

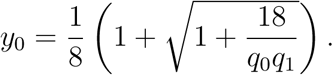

For the threshold *y*_0_ to be smaller than 1, we need that *q*_0_*q*_1_ ≥ 3/8. If app users are 100% reliable, that is *q*_0_*q*_1_ = 1, we get that the minimal adoption rate of the app is

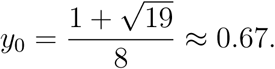

Let us summarize this part:

- to get from effective *R*_0_ = 3/2 to *R*_0_ = 1, the minimal adoption rate of the app assuming perfect cooperative behavior is *y*_0_ = 77%.
- to get from effective *R*_0_ = 4/3 to *R*_0_ = 1, the minimal adoption rate of the app assuming perfect cooperative behavior is *y*0 = 67%.

Figure 5 shows more generally how *y*_0_ varies as a function of *R*_0_ for different parameter values of *f* and *q*_1_. The previous numerical results can be pinpointed on top right panel of Figure 5 (corresponding to *f* = 1/3 and *q*_1_ = 1).

**Figure 5.**
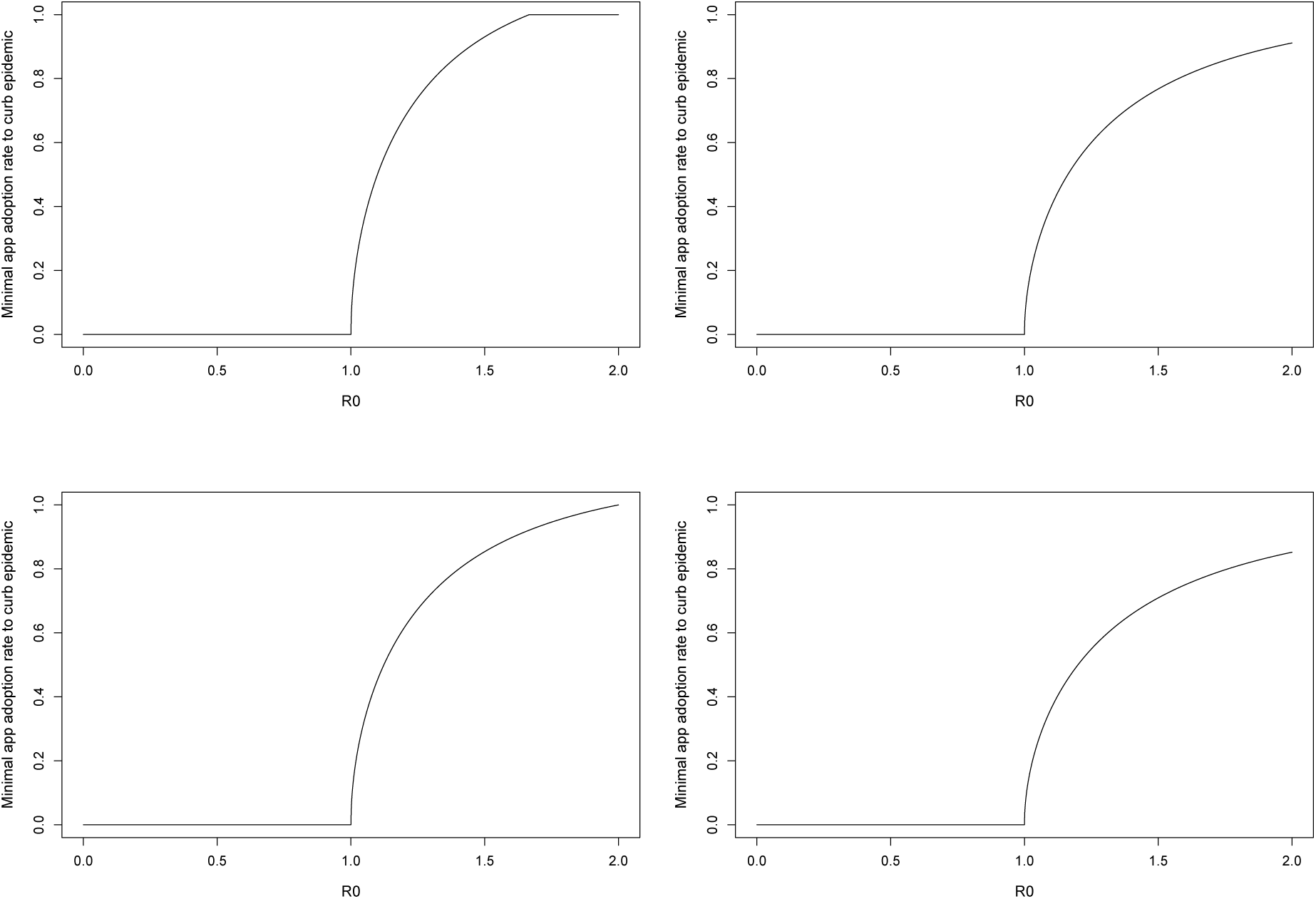
Minimal app adoption rate *y*_0_ to curb epidemic as a function of effective *R*_0_ under forward contact tracing. Rows differ according to fraction *f* of asymptomatics and columns differ according to probability *q*_1_ of cooperating (i.e., of quarantining upon app alert). The probability of informing app upon symptoms is *q*_0_ = 1. When forward tracing is unable to curb epidemic, *y*_0_ is set equal to 1 by convention (top left panel). **First row:** *f* = 1/3. **Second row:** *f* = 1/6. **First column:** *q*_1_ = 0.6. **Second column:** *q*_1_ = 1.

It can be seen in Figure 5 that the minimal adoption rate of the app to reduce *R*_0_ below 1 increases very steeply from *R*_0_ = 1, requiring the fraction *y* of app users to be very large, even in the best scenarios when the fraction *f* of asymptomatics is small and the probability *q*_1_ of cooperation is large (bottom right). There is little chance that (at least, Western) countries can access to the required rates of adoption, unless social distancing already curbs *R*_0_ to values very close to 1.

We will now study the case of **recursive contact tracing**.

## 5.1 Recursive contact tracing

### Preliminary observations

In this section, we assume that alerts can be of degree 2 in the graph of contacts. Since the transmission tree is a subgraph of the contact network (assumed to also be tree-like), a physical contact of degree 2 is either a sibling or a grand-daughter in the transmission tree.

If a contact of degree 1 or 2 related to an index case decides to cooperate and self-quarantine upon being alerted, we will assume that this quarantine:

1. removes her from the epidemic if she is a daughter of the index case (contact of degree 1, alerted by her mother), as previously;
2. has no effect on her if she is the mother of the index case (contact of degree 1, alerted by her daughter), as previously;
3. **removes her from the epidemic if she is a grand-daughter** of the index case (contact of degree 2, alerted by her mother);
4. **removes her from the epidemic with probability** *ℓ* **if she is a sibling** of the index case (contact of degree 2, alerted by her mother): the idea is that she is removed iff she is infected by their common mother later (or say, at least two days later) than her sib, which occurs with probability *ℓ* (default value *ℓ* = 1/2).

In the next subsection, we quantify the effect of the latter type of removals as listed previously.

### Quantifying the effect of sibling’s alerts

In this section, we take into account the fact that a *Y*-individual with a *Y*-mother may be alerted by her mother as contact of degree 2 of her siblings of type *Y I*.

Assume that the mother is of type *Y Z*, *Z* = *A* or *I*. We denote by **t_z_** the **probability that an individual of type** *Y***, daughter of an individual of type** *Y Z***, does not comply to any alert coming from her siblings**. We also denote by **x_z_** the **probability of not being removed despite these alerts**. Because we have assumed that only a fraction *ℓ* of compliant individuals (alerted by their sibling via their common mother) are removed (interpreted as: ‘infected later than alerting sibling’), we have

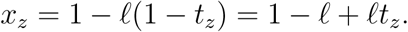

**Figure 6.**
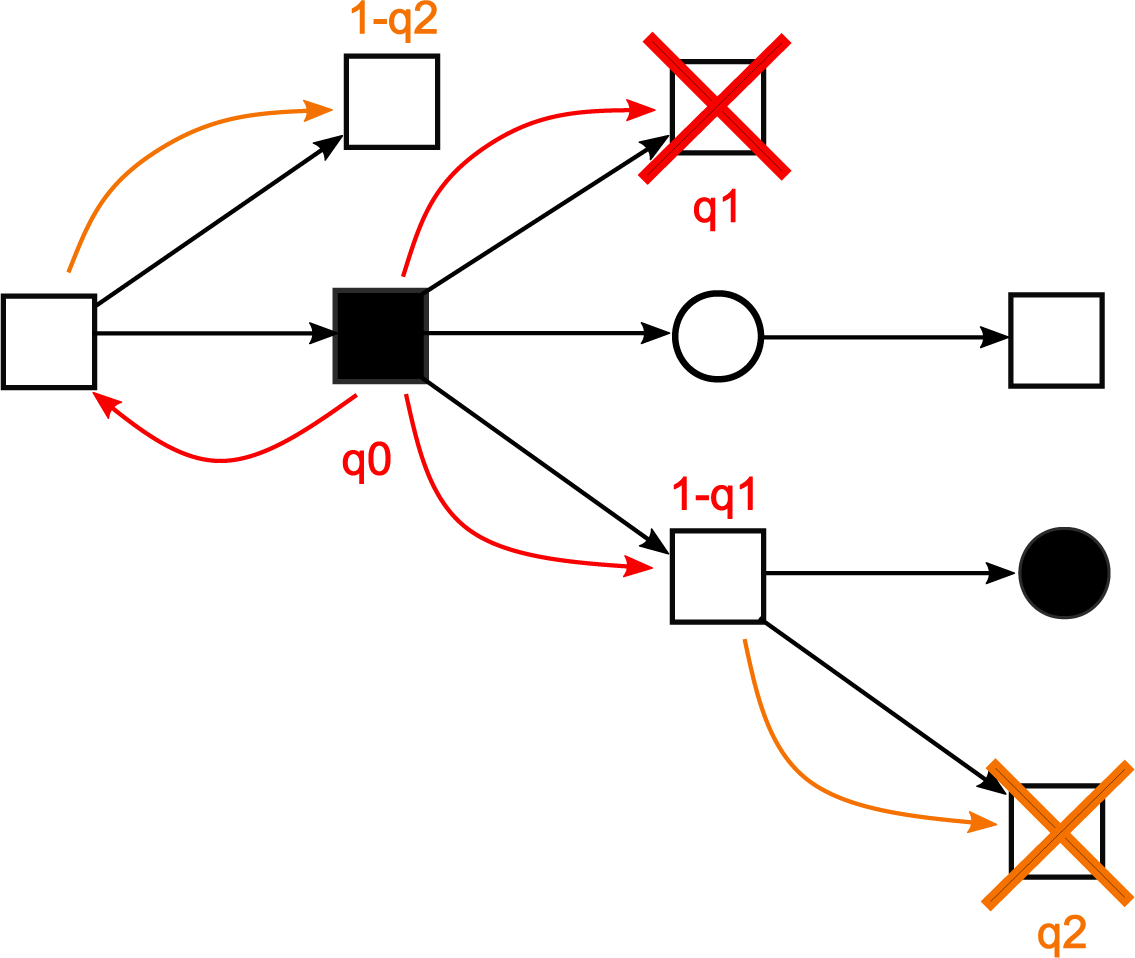
A *Y I*-individual (symptomatic, using the app) decides with probability *q*_0_ to inform the app of her symptoms. This results in alerting all her physical contacts (degree 1, red arrows) of type *Y* (among which her mother and daughters in the epidemic) and in alerting all the contacts (degree 2, orange arrows) of type *Y* of these contacts (among which her siblings and grand-daughters in the epidemic). Each alerted daughter (degree 1) does self-quarantine with probability *q*_1_ and is then removed from the epidemic (red cross). Each alerted grand-daughter (degree 2) does self-quarantine with probability *q*_2_ and is then removed from the epidemic (orange cross). Each alerted sibling (degree 2) does self-quarantine with probability *q*_2_ and is then removed from the epidemic with probability *ℓ* (not shown). Legend: white = **A**, black = **I**, square = **Y**, circle = **N**.

Now let us express *t_z_* thanks to the parameters of the model:

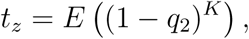

where *K* is the number of *Y I* cooperative individuals among the *N_z_* − 1 siblings of the focal individual. This yields

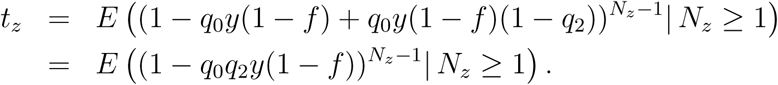

In applications we will assume that *N_z_* follows the Poisson distribution with parameter *R_z_*, so that

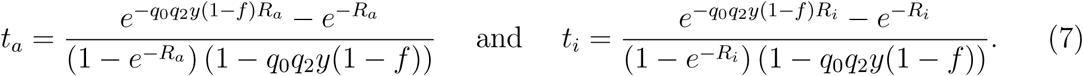

As soon as *q*_0_, *q*_2_ or *y* is zero, we find as expected that *t_z_* = 1 and so *x_z_* = 1.

### Stopping line technique

In this section, we prove Theorem 1(ii).

**Figure 7.**
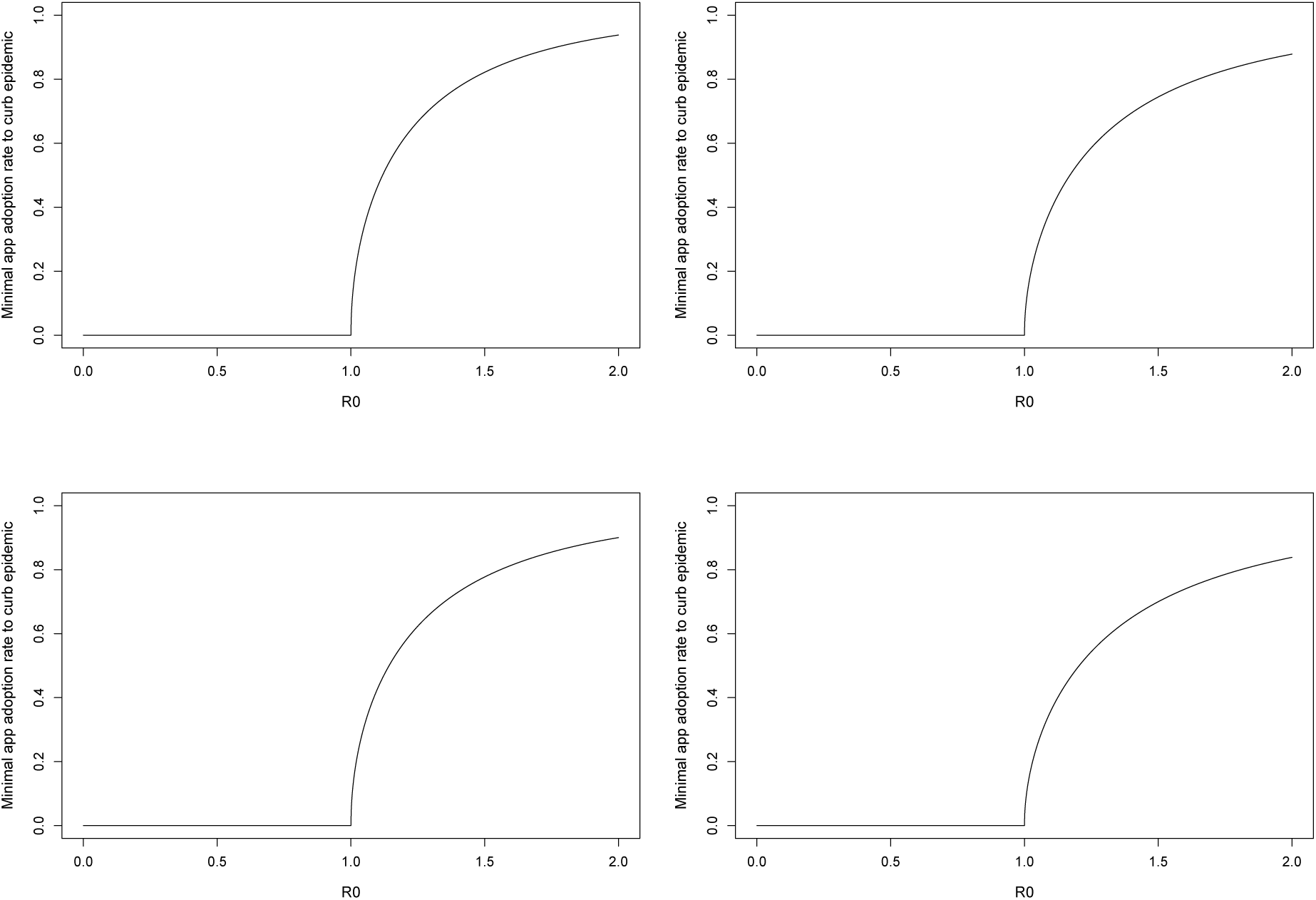
**Minimal app adoption rate** *y*_0_ **to curb epidemic as a function of effective** *R*_0_ **under recursive contact tracing**. Rows differ according to fraction *f* of asymptomatics and columns differ according to probabilities *q*_1_ and *q*_2_ of cooperating (i.e., of quarantining upon app alert as a contact of degree 1/of degree 2). The probability of informing app upon symptoms is *q*_0_ = 1. **First row:** *f* = 1/3. **Second row:** *f* = 1/6. **First column:** q_1_ = *q*_2_ = 0.6. **Second column:** *q*_1_ = *q*_2_ = 1.

Because the behavior of an individual who is alerted possibly influences the removal of her daughters, we have to distinguish whether a *Y*-individual has received an alert or not. A *Y*-individual who has been alerted will be said ‘in excited state’ or simply ‘alerted’ and the corresponding type denoted with a star. Here are the following kinds of types to consider: *Y I*\*, *Y A\**, *Y I*, *Y A*, *N A*, *N I*. An individual of the four latter types will be said ‘in ground state’.

An individual in ground state who is in state *Y A* or *Y I* (resp. *N A* or *N I*) with probabilities *f* and 1 − *f* will merely be denoted *Y* (resp. *N*) and called a **regenerative state**. In the genealogical tree of transmissions starting from a single individual, **we follow all lines of descent descending from her and stop them at the first regenerative state** encountered. The set of regenerative states forms what is called a **stopping line** [2] in the transmission tree. We will call **seed-tree** the tree obtained by pruning from the initial transmission tree all vertices downstream of the stopping line. The leaves of a seed tree are all in a regenerative state, either *Y* or *N*. We call them the *Y***-regenerative leaves and** *N***-regenerative leaves** of the seed tree, respectively.

We can then define a **Galton–Watson branching process with two types** *Y* and *N* by saying that **the offspring of type** *Z* (*Z* = *Y* **or** *N*) **of a** *X***-individual** (*X* = *Y* **or** *N*) **are the** *Z***-regenerative leaves of a seed-tree seeded by** *X*. This process has no interest in itself except that the epidemic dies out iff it is subcritical.

We let *m_ya_,_y_* denote the average number of *Y*-regenerative leaves of a seed tree seeded by a *Y A*-individual. We define similarly *m_yi_,_y_, m_ya_,_n_, m_yi_,_n_, m_na_,_y_, m_ni_,_y_, m_na_,_n_, m_ni_,_n_*. We then define

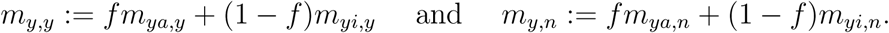

Similarly, we define

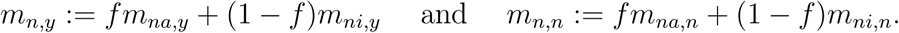

We still define *M* as the matrix with entries *m_y_,_y_, m_y_,_n_, m_n_,_y_* and *m_n_,_n_*, despite the fact that these quantities have a different meaning from theirs in the previous section. However, **since** *M* **is the mean matrix of the two-type branching process defined previously, we still have that the epidemic dies out iff the leading eigenvalue of** *M* **is smaller than 1**.

We will now compute the expected number of individuals of each type at generation k of a seed-tree, for example denoted [*Y I*\*]*_k_* for individuals of type *Y I*\*. Let us make some preliminary observations:

- A seed-tree seeded by a *N A* a *N I* or a *Y A*-individual stops at generation 1, because all her daughters are in ground state (*Y* or *N*).
- When the seed-tree is seeded by a *Y I*-individual, there are two possibilities:
  –if the seed cooperates (i.e., informs the app), then the daughters of the seed can be of type *Y A*\*, *Y I\** or *N*;
  –if the seed does not cooperate, the seed-tree stops at generation 1 as previously.

Now we consider a seed-tree starting from a *Y I*-individual. Recall that for the infection of a multiply alerted individual to succeed, this individual must defect independently to all alerts she has received.

#### Infections from an individual of type *Y I* of generation 0 (seed)

With probability 1 − *q*_0_, the daughters of the seed are

- an expected number *yx_i_ R_i_* of type *Y*,
- an expected number (1 − *y*)*R_i_* of type *N*.

Recall that 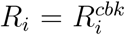, where the values of *c, b, k* and *r* can be tuned once for all. With probability *q*_0_, the daughters of the seed are

- an expected number *y f* (1 − *q*_1_)*x_i_R_i_* of type *Y A\**,
- an expected number *y*(1 − *f*)(1 − *q*_1_)*x_i_R_i_* of type *Y I\**,
- an expected number (1 − *y*)*R_i_* of type *N*.

**Figure 8.**
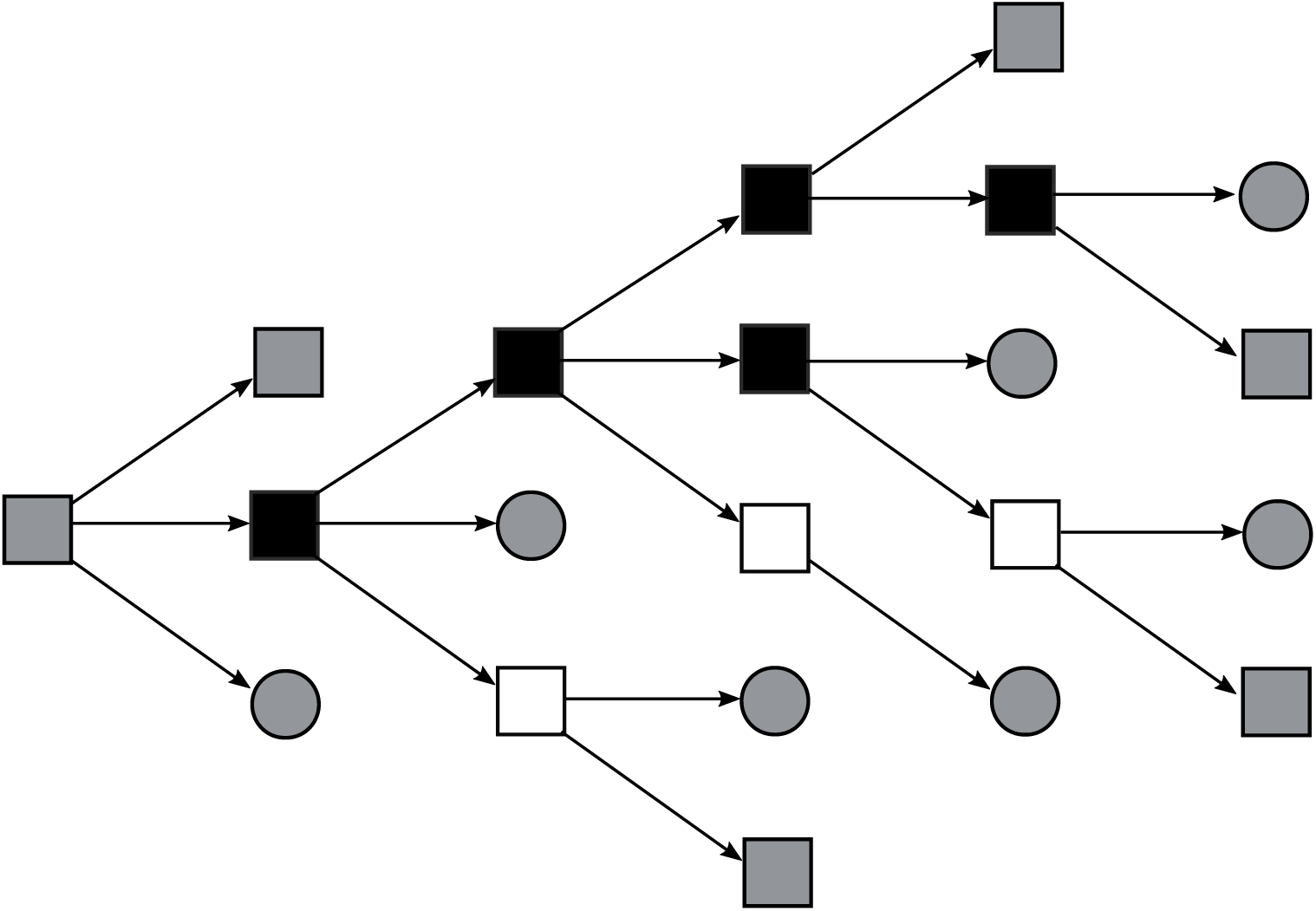
A seed-tree starting from a *Y*-individual. A seed-tree is a maximal subtree of the transmission tree such that all **internal** transmission edges are doubled with an alerting arrow (not shown). In particular, a leaf of a seed-tree is by definition an individual who receives no alert (also called regenerative), either because she is of type *N* or because her mother is a *Y I* who does not cooperate, or because her mother is a *Y A* merely forwarding an alert (degree 2). Legend: white = **A**, black = **I**, square = **Y**, circle = **N**, gray = **regenerative state**.

#### Infections from an individual of type *Y A\** of generation *k* ≥ 1

Daughters of an individual of type *Y A\** of generation *k* ≥ 1 are

- an expected number *y*(1 − *q*_2_)*x_a_R_a_* of type *Y*,
- an expected number (1 − *y*)*R_a_* of type *N*.

#### Infections from an individual of type *Y I\** of generation *k* ≥ 1

With probability *q*_0_, daughters of an individual of type *Y I\** of generation *k* ≥ 1 are

- an expected number *y f*(1 − *q*_1_)(1 − *q*_2_)*x_i_R_i_* of type *Y A\**,
- an expected number *y*(1 − *f*)(1 − *q*_1_)(1 − *q*_2_)*x_i_R_i_* of type *Y I\**,
- an expected number (1 − *y*)*R_i_* of type *N*.

With probability 1 − *q*_0_, her daughters are

- an expected number *y*(1 − *q*_2_)*x_i_R_i_* of type *Y*,
- an expected number (1 − *y*)*R_i_* of type *N*.

Then we obtain the following equations

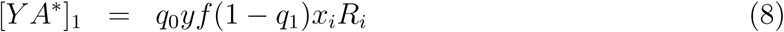

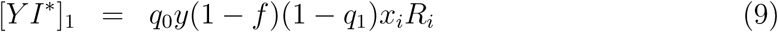

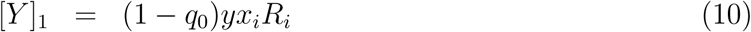

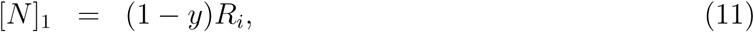

and for any *k* ≥ 1,

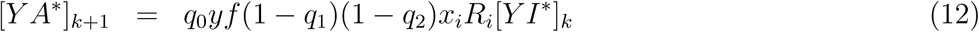

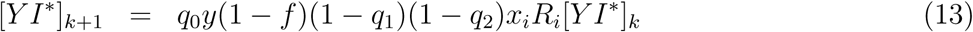

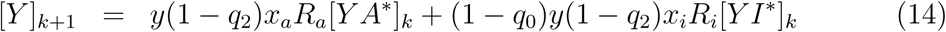

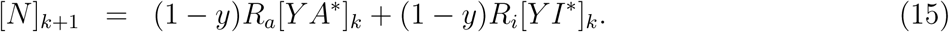

Now define *T_y_* (resp. *T_n_*) the total expected number of *Y*-regenerative (resp. *N*- regenerative) leaves of the seed-tree seeded by a *Y I*-individual:

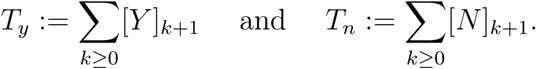

First observe that thanks to (9) and (13), we get

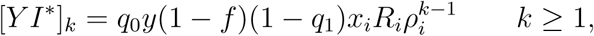

with

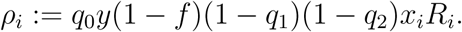

As a consequence, thanks to (8) and (12),

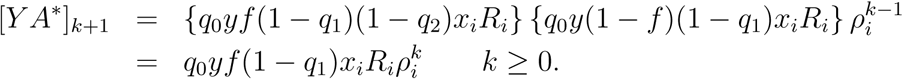

Next, thanks to (14), we get

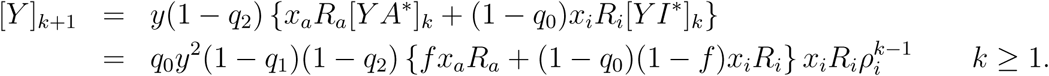

Finally, thanks to (15),

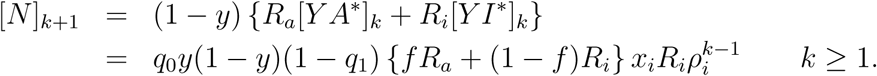

Using (10), we have

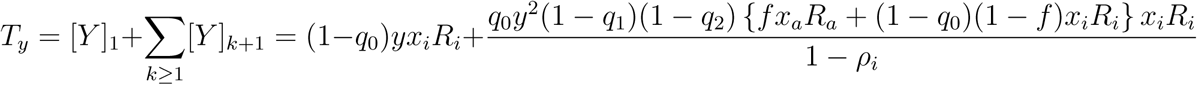

Similarly, using (11), we have

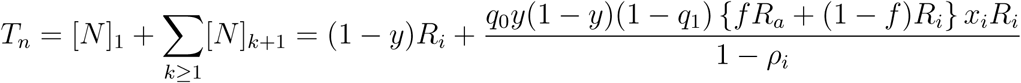

Now we use the fact that

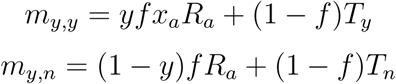

while

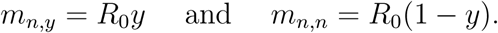

Elementary algebra yields

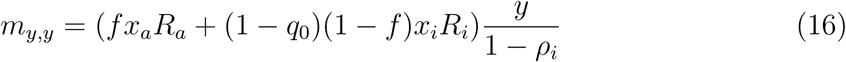

and

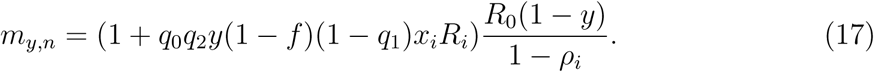

The determinant of *M* is

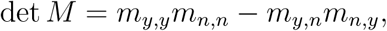

which after calculation is

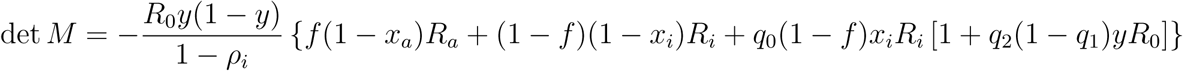

Also recall that the trace of *M* is Tr *M* = *m_y_,_y_* + *m_n_,_n_*. Now as in the case of forward tracing, we denote by *Q* the characteristic polynomial of *M*, i.e.,

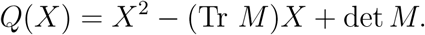

Again *Q*(0) ≤ 0 so the leading eigenvalue of *M* is the unique positive root of *Q*. Also, this root is smaller than 1 if and only if *Q*(1) ≥ 0, which yields

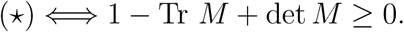

After some algebra, we get

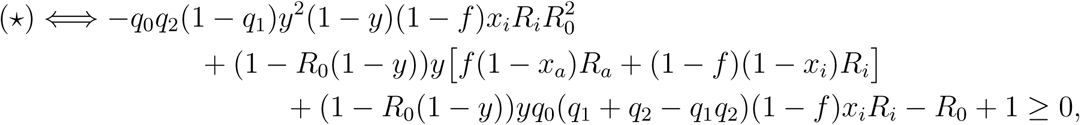

which is exactly Eq (5) and so ends the proof.

## 6 Discussion

### Robustness of results

Contact tracing supposes that case isolation is possible and is in force. Since contact tracing has no effect (at least in democracies?) on the efficiency of case isolation, and case isolation can be enforced independently of contact tracing, a measure of the effect of contact tracing should not include the effect of case isolation. This explains why we have chosen to express the minimal rate *y*_0_ (of adoption of a contact tracing app to curb the epidemic) as a function of the effective *R*_0_, that is, the *R*_0_ obtained by non-electronic interventions, notably case isolation.

By measuring only the net effect of contact tracing, our results are effectively insensitive to assumptions on the natural value of *R*_0_, as well as on crucial parameters like *f* (fraction of asymptomatics) and *b* (roughly speaking, fraction of infections made before symptoms). We also found empirically that our results hold also for a wide range of values of the cooperating probabilities, as testified by the striking similarity of the four panels of Figure 7.

### Interpretation of parameters

Let us discuss briefly the interpretation of some parameters in terms of the natural history of the virus and of the nature of healthcare policies.

- *b* is the fraction of secondary infections barred thanks to case isolation. Its value depends both on the time *T* taken to actually self-isolate after day *D* of symptom onset and on the natural history of the virus, via the fraction *m* of secondary infections made before *D* + *T*. We have parameterized *b* as *b* = 1−*p*+*pm*, where *p* is the probability of actually self-isolating. Parameter *b* can be tuned optimally by minimizing *T* so as to minimize *m* and by testing more systematically so as to maximize *p*: a symptomatic individual tested positive may feel more inclined to self-isolate.
- *q*_0_ is the probability that a symptomatic app user informs the app of her symptoms when they first appear. In applications, we have assumed throughout the manuscript that *q*_0_ = 1, but we cannot discard the existence of individuals who will download the app only to be aware of whether they have been in contact with sick individuals (leecher vs seeder).
- *q*_1_ (resp. *q*_2_) is defined here as the probability of self-quarantining upon being alerted by app from a contact of degree 1 (resp. of degree 2). In this sense, we have *q*_1_ = *q*_2_ whenever app users are not aware of whether they are contacts of degree 1 or 2 of the alerting index case; if they are, it is more reasonable to assume *q*_2_ < *q*_1_. Again, testing more systematically can help increase *q*_1_ and *q*_2_. Alternatively, the quantity *q*_1_ (resp. *q*_2_) can be interpreted as the likelihood of actual removal from the epidemic as a daughter of index case (resp. as a grand-daughter of index case). In this interpretation, one has on the contrary *q*_1_ < *q*_2_, bearing in mind that some daughters are infected too early to be actually removed, as opposed to grand-daughters.

### Comparing forward and recursive contact tracing

The effect of recursive tracing is two-fold: to secure removal of grand-daughters of index cases when removal of daughters has failed (*q*_1_ < 1, see Remark 2) and to remove siblings of index cases by alerting their mother (see Remark 3). Note that secondary contacts of an index case combine an average number 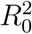 of grand-daughters and an unknown, larger number of non-infected physical contacts (of physical contacts). Due to the psychological and economical costs of quarantining all these secondary contacts, it is important to evaluate the marginal benefit of recursive tracing compared to forward tracing.

Figure 9 compares the minimal app adoption rate required to curb the epidemic with forward (1st row) vs recursive (2nd row) tracing, when *q*_1_ = 0.6 (1st column) and when *q*_1_ = 1 (2nd column). When *q*_1_ = 0.6, alerts of degree 2 (recursive tracing) can rescue the failure of forward tracing to curb the epidemic when the effective *R*_0_ is high, provided *y* is accordingly high. When *q*_1_ = 1, the only benefit of recursive tracing is through alerting siblings, and we see by comparing top right and bottom right panels that this benefit is hardly detectable. In addition, whatever the value of *q*_1_, both strategies have basically the same effect for small values of *R*_0_ and *y*.

**Figure 9.**
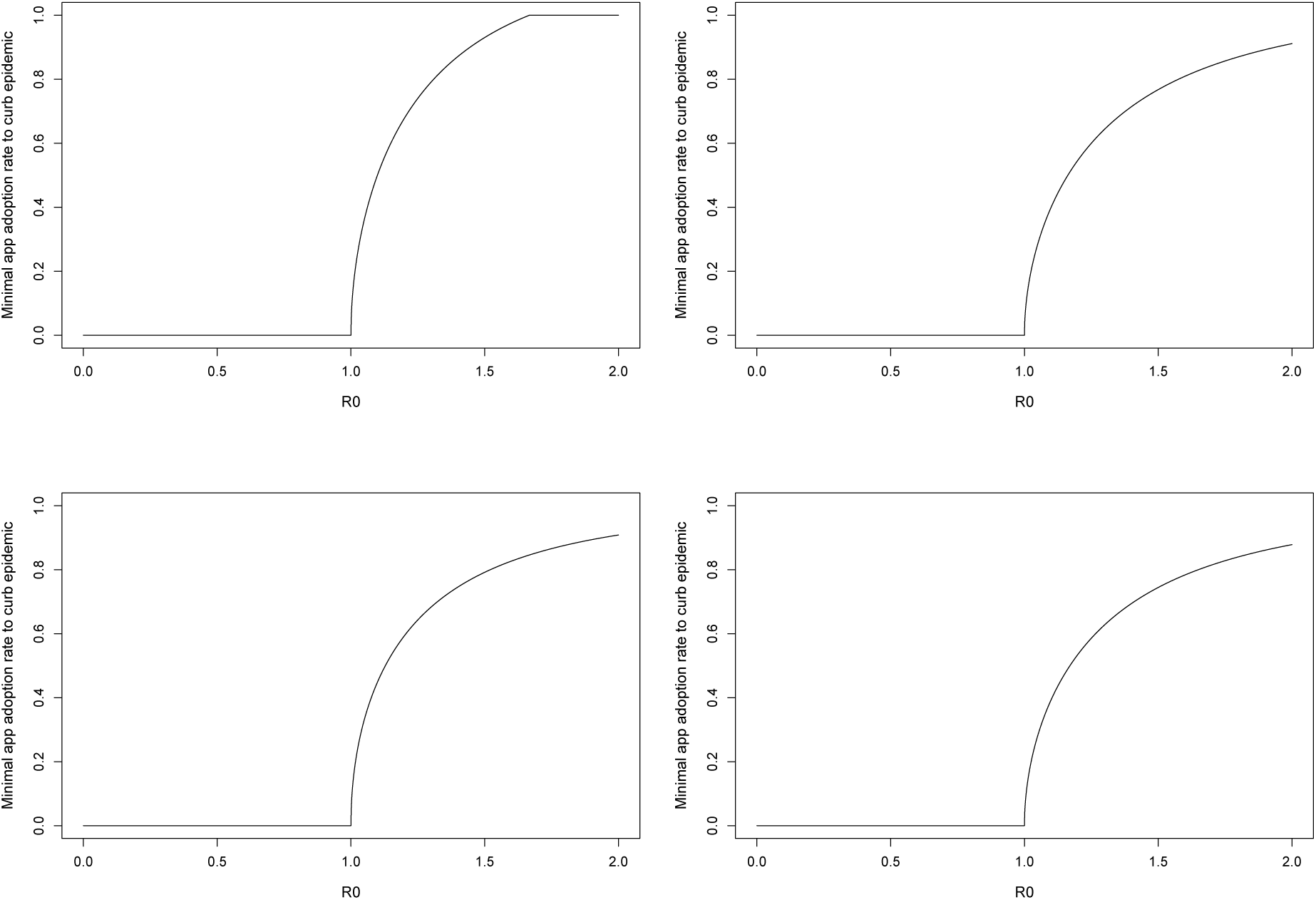
Minimal app adoption rate *y*_0_ to curb epidemic as a function of effective *R*_0_ under forward vs recursive contact tracing. \\\\Rows differ according to mode of contact tracing and columns differ according to probability *q*_1_ = *q*_2_ of cooperating (i.e., of quarantining upon app alert). The probability of informing app upon symptoms is *q*_0_ = 1 and the fraction of asymptomatics is *f* = 1/3. In the case of recursive tracing, the probability of quarantining is *q*_2_ = 1. When forward tracing is unable to curb epidemic, *y*_0_ is set equal to 1 by convention (top left panel). **First row:** Forward tracing. **Second row:** Recursive tracing. **First column:** *q*_1_ = 0.6. **Second column:** *q*_1_ = 1.

We conclude that the marginal benefits of recursive tracing are negligible compared to its costs, so that in particular, the explicit result given in Eq (4) can be used for all practical purposes.

### Relation to previous work

The model (but not the approach) that we use here is similar to the one used in [6] and in two other works specifically interested in the current epidemic [5, 3]. See also [4, 13] for seminal works on the topic of quantifying the effect of non-pharmaceutical interventions on epidemics and [15, 17] for works on contact networks and contact tracing.

We now explain why our predictions seem somewhat less optimistic than those given in [5, 3]. In these works, the *R*_0_ given corresponds to what we have termed 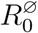 or 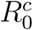 possibly taking into account social distancing (in view of the values considered) but not case isolation. Recall that case isolation is parameterized in our notation by the fraction *b* of infections made before isolation. Also recall that with our notation, symptoms appear *D* days after infection, case isolation occurs *T* days after symptoms and m is the fraction of infections made before *D* + *T*, so that *b* = 1 − *p* + *pm*, where *p* is the probability of actually self-isolating.

In [5], the baseline scenario has *f* = 0 (or 0.1), *T* = 3.4 (‘short delay’) and the fraction of infections made before *D* is 0.15, which corresponds to *b* ≈ 0.7 (see Figure 2 in this paper). In addition, the only scenario for which contact tracing works has 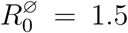. This corresponds to an effective *R*_0_ equal to 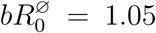 when *f* = 0 (and equal to 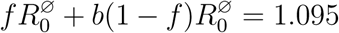 when *f* = 0.1), in agreement with our findings that moderate adoption rates of the contact tracing app are sufficient only when the effective *R*_0_ is very close to 1.

In [3], 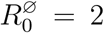 and 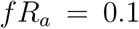. The main results can be seen on Figure 3 in this paper. Each panel corresponds to a different value of *T*, decreasing from left to right. The rightmost panel (best case scenario) has *T* = 0, which implies that m is the fraction of infections made before symptoms by symptomatics and is approximately 0.5 (see Figure 2 in this paper). The panel shows the region of parameter space (*X*, *Y*) for which the epidemic dies out, where *X* is the ‘success rate of instant isolation of symptomatic cases’ and *Y* is the ‘success rate of instant contact tracing’. In our notation, *X* = *p* and *Y* = *q*_0_*q*_1_*y*^2^. The effective *R*_0_ is roughly 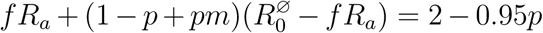, ranging from 1.05 to 2 when *p* ranges between 0 and 1. Taking *q*_0_ = *q*_1_ = 1 and referring to top right panel of our Figure 5, our prediction is that *y*_0_ ranges between 0.2 and 0.9 as *p* goes from 1 to 0, i.e. 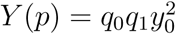 ranges between 0.04 to 0.8, which is actually slightly more optimistic for large values of p than what shows Figure 3 in [3]. Referring to calculations made page 12 and taking *p* = 1/2 so that the effective *R*_0_ ≈ 3/2, our prediction is *y*_0_ ≈ 0.77. This yields 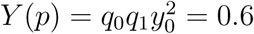, which is visually the same prediction as on Figure 3 in [3].

In conclusion, we see that our predictions are actually in line with those given in [5] and [3]. The most prominent differences come from the facts that 1) we measure the net effect of contact tracing by comparison with an effective *R*_0_ that takes into account the effect of case isolation (as a rule of thumb there can be a factor 2 between the effective *R*_0_ and the natural *R*_0_ other studies refer to) and 2) we measure this effect in terms of a minimal rate of app users rather than in terms of a minimal efficiency of contact tracing (as a rule of thumb the latter is the square of the former). The bottomline is that all three studies agree that the minimal rate of contact tracing app users must be larger than 60-70% to curb the epidemic unless the effective *R*_0_, already taking case isolation and social distancing into account, is already very close to 1.

## Data Availability

No data

## Acknowledgements

The author thanks the *Center for Interdisciplinary Research in Biology* (CIRB) for funding and his entire research group SMILE (Stochastic models for the inference of life evolution) for numerous fruitful (online) discussions around the COVID-19 pandemic.

